# Is there a spinal tap responder in progressive supranuclear palsy?: The first prospective study

**DOI:** 10.1101/2023.09.28.23296229

**Authors:** Masahiro Ohara, Takaaki Hattori, Qingmeng Chen, Kaoru Shimano, Kosei Hirata, Mie Matsui, Takanori Yokota

## Abstract

**Objective:** Progressive supranuclear palsy (PSP) is a progressive neurodegenerative disease, and sometimes shows idiopathic normal pressure hydrocephalus (iNPH)-like presentations. We aimed to evaluate spinal tap responsiveness in patients with PSP, including the effect of sham spinal tap.

**Methods:** Eleven patients with PSP, ten with probable/definite iNPH, and eight control patients were prospectively enrolled. All participants underwent sham spinal tap and spinal tap procedures. Gait was evaluated using wearable inertial sensors. We defined “tap responders” as individuals with a 10% or more improvement from baseline in any of the gait parameters (timed up-and-go test total time, stride length, and velocity during straight walking under single-task and cognitive dual-task conditions). We compared the ratio of tap responders in patients with PSP to patients with iNPH and controls.

**Results:** The ratio of tap responders in patients with PSP was significantly higher than that in control patients, but not different from patients with iNPH. The ratio of sham tap responders was significantly higher in PSP patients than that in control patients, but not that in patients with iNPH. Notably, one patient with PSP responded to the spinal tap beyond the effect of sham spinal tap, and even to the shunt operation.

**Interpretation:** This is the first prospective study to demonstrate tap and shunt responsiveness in patients with PSP while highlighting the placebo effects of the spinal tap in patients with PSP and iNPH. Our findings suggest that some PSP patients have impaired cerebrospinal fluid circulation, contributing to a distinct component of the clinical spectrum.

## Introduction

Progressive supranuclear palsy (PSP) is a progressive neurodegenerative disease that is characterized by supranuclear gaze palsy, akinetic rigidity, gait disturbance, and dementia.^1^ PSP presents with various clinical phenotypes that mimic other diseases^2^ and occasionally manifests with idiopathic normal pressure hydrocephalus (iNPH)-like presentations.^3,4^ iNPH is a clinical disease entity that is characterized by the triad of gait disturbance, dementia, and urinary disturbance, and these symptoms can be improved by shunt operation.^5^ We previously demonstrated that PSP develops iNPH-like MRI features, particularly a high disproportionately enlarged subarachnoid space hydrocephalus (DESH) score (≥ 5), more often than other neurodegenerative diseases, and proposed a concept of PSP with hydrocephalus (PSP-H).^4^ Similarly, the concept of NPH secondary to neurodegenerative diseases (neurodegenerative NPH) has been also proposed^6^, and PSP is one of the most common etiologies of neurodegenerative NPH.^3^ However, no prospective studies have evaluated iNPH-like clinical presentations, including spinal tap/shunt responsiveness, in patients with PSP. Previous literature has reported autopsy-proven patients with PSP who responded to the shunt operation with an antemortem diagnosis of iNPH,^3,6–9^ indicating the existence of iNPH-like clinical features in patients with PSP. We hypothesize that some symptoms of PSP would be improved by spinal tap/shunt operation albeit temporarily, based on these reports.

Spinal tap responsiveness is a distinct feature of iNPH and is thus included in the diagnostic criteria for probable iNPH in the Japanese guideline.^10^ However, a conventional spinal tap presents several challenges for accurately diagnosing iNPH. The potential placebo effects of spinal taps and shunt operations on the symptoms have been discussed.^11^ A review study performed by Espay et al. showed that the response rates to the shunt operation in patients with iNPH varied from 31% to 89%.^6^ This variable response rate to the shunt operation indicates that the shunt operation is often performed for inappropriate individuals, including those who seemed to “respond” to the spinal tap by a placebo effect.^11^

Furthermore, the spinal tap exhibits an average specificity of 75% in predicting the outcomes of shunt operation,^12^ thereby emphasizing the issue of false positives associated with this diagnostic approach. A case report has documented the utility of a sham spinal tap to address the challenge of low diagnostic accuracy.^13^ Therefore, in order to ascertain the existence of patients with PSP having spinal tap responsiveness, we employed a novel protocol that incorporated both a spinal tap and a sham spinal tap.

In this study, we aimed to evaluate the effect of spinal taps and sham spinal taps on gait and cognitive function in patients with PSP compared to those with iNPH or control patients.

## Material and methods

### Participants

Patients with PSP and participants with hydrocephalus, as defined by a high Evans index > 0.30,^14^ were prospectively recruited at an outpatient clinic in the Department of Neurology at Tokyo Medical and Dental University Hospital from 2018 to 2022. The clinical diagnosis was made according to the Movement Disorder Society Clinical Diagnostic Criteria for clinically suggestive of, possible, or probable PSP^15^ and the Japanese guidelines for clinically probable or definite iNPH.^10^ Although the criteria for PSP requires the exclusion of the presence of hydrocephalus, the diagnosis of PSP was made when all other diagnostic criteria for PSP were fulfilled even if hydrocephalus was present since the concept of PSP-hydrocephalus (PSP-H)^4^ or neurodegenerative NPH^6^ has been proposed. Patients were included if they were older than 40 years old at enrollment and had no current medical history with anti-thrombotic medications. Patients with brain tumors, intracranial hemorrhage, bleeding tendency, difficulties in walking for at least 20 meters, or skin infection at the lumbar puncture site were excluded.

### Study design

All patients underwent gait analysis five times: twice within 5 days before the sham spinal tap, once within 1 day after the sham spinal tap, and twice within 5 days after the spinal tap. Better gait parameters obtained before the sham spinal tap and after the spinal tap were used for analysis. Additional gait analysis was conducted following the operation in patients who underwent a shunt operation.

Neuropsychological tests were performed within 7 days before and 3 days after the spinal tap. In addition to gait analysis and neuropsychological tests, baseline disease severity was evaluated using the Motor Examination of Movement Disorder Society Unified Parkinson’s Disease Rating Scale (MDS-UPDRS part III)^16^ and the iNPH grading scale (iNPHGS).^17^ Motor function, including gait, was assessed in all patients by a board-certified neurologist (M.O.). The Institutional Review Boards of Tokyo Medical and Dental University Hospital approved this study. All data were collected as a part of a prospective study and written informed consent was obtained from all participants.

### Brain MRI and DAT- and MIBG-SPECT

The Evans index,^14^ the callosal angle in the coronal plane at the posterior commissure,^18^ the callosal angle at the splenium^19^ and DESH score^20^ were evaluated using 3-dimensional T1-weighted image to evaluate the iNPH-like MRI features. A high DESH score (≥ 5) is a useful predictor of positive responsiveness to shunt operation in patients with iNPH^20^, and may also be useful in characterizing iNPH-like MRI features in neurodegenerative disorders including PSP.^4^ To ascertain the pathological background, all patients underwent dopamine transporter- and cardiac ^123^I-metaiodobenzylguanidine-single photon emission computed tomography (DAT-SPECT and cardiac MIBG-SPECT, respectively). Abnormalities on DAT-SPECT were assessed by visual evaluation^21,22^ and quantitative analysis.^23^ We employed DaTView (Nihon Medi-Physics Co., Ltd.) which compares the DAT uptake value of the basal ganglia to the mean DAT uptake value of the whole brain excluding the basal ganglia. We compared patients’ specific binding ratio with that of the age-matched healthy individuals to quantify dopaminergic denervation. The abnormality of cardiac MIBG-SPECT was evaluated based on the low early heart/mediastinum (H/M) ratio^24^ or delayed H/M ratio,^24,25^ compared with the standardized cut-off values of healthy controls. Cardiac MIBG-SPECT exhibits high sensitivity and specificity (approximately 90%) in distinguishing Lewy body diseases and disease controls.^26^

### Gait analysis

The participants’ gait was measured and analyzed using inertial measurement units (IMUs; WALK-MATE Viewer^®^, WALK-MATE LAB., Tokyo, Japan) attached by bilateral bands at the ankles and hips.^27^ All patients were instructed to walk fast-paced to minimize inter-trial variation. Stride length and stride velocity were measured under the following two conditions: 1) walking a distance of 15 m (straight walking) and 2) straight walking under a cognitive dual-task (walking with serial 7 subtractions from 500). In addition to straight walking, the timed up-and-go test (TUG) was performed using a standardized method.^28^ The patients performed the TUG 3 times; the mean TUG total time was used for analysis. Tap responders to gait function were defined by those who exhibited a 10% improvement from baseline^29^ for any of the gait parameters (TUG total time, stride length, and stride velocity during straight walking under single and cognitive dual tasks).

### Neuropsychological tests

Global cognition was first evaluated by using the Japanese version of the Montreal Cognitive Assessment test (MoCA-J).^30^ Then, comprehensive neurocognitive assessments, which were composed of six cognitive domains, were performed by neuropsychologists. Six cognitive domains were evaluated by at least two batteries for each cognitive domain as follows: 1) Immediate memory: list memory and episode memory (subsection of the Japanese version of the repeatable battery for the assessment of neuropsychological status (RBANS)^31^), 2) Delayed memory: list recall, list recognition, episode recall, and figure recall (subsection of RBANS), 3) Visuospatial function: figure copy and line orientation (subsection of RBANS), 4) Language: picture-naming and category fluency (subsection of RBANS) as well as auditory verbal comprehension (subsection of Western Aphasia Battery^32^), 5) Attention/working memory: digit span and letter-number sequencing subtests (subsection of RBANS), and 6) Executive function: the rule shift cards test (subsection of the Behavioral Assessment of the Dysexecutive Syndrome^33^), letter fluency test^34^ and time to complete part A and part B of trail-making test.^35^ Each test score was converted to a Z-score by using age-matched norms. In a previous report about the diagnostic criteria for mild cognitive impairment in Parkinson’s disease, deficits of at least −1.5 standard deviations (SD) on two or more subtests in neuropsychological tests were reported to be suitable to diagnose Parkinson’s disease with mild cognitive impairment.^36,37^ Based on these reports, tap responders to cognitive function were defined as those who exhibited a 1.5 or more improvement in the Z-score from baseline in at least two or three cognitive batteries.

### Outcome measures

The primary outcome was to evaluate the inter-group difference between patients with PSP and iNPH or control patients regarding the ratio of the spinal tap responders to gait function. The secondary outcome was to examine the inter-group differences between patients with PSP and iNPH or control patients in the ratio of the spinal tap responders to cognitive function and in the ratio of the sham spinal tap responders to gait function.

### Statistical analyses

Statistical analyses were performed using the GraphPad Prism 9 software (GraphPad Software, San Diego, CA, USA). For continuous variables, inter-group differences between patients with PSP and the other groups were evaluated using the Kruskal-Wallis test followed by Dunn’s test. For categorical variables, inter-group differences between patients with PSP and the other groups were evaluated by using the chi-square test followed by Fisher’s exact test with the Bonferroni correction. The Friedman test followed by Dunn’s test was performed to analyze the changes in gait parameters in the three groups over time (at baseline, after the sham spinal tap, and after the spinal tap),

## Results

### Demographic and clinical data in the three groups

Eleven drug-naïve patients with PSP (nine with probable PSP with Richardson’s syndrome (PSP-RS) and two with suggestive PSP with predominant postural instability) and 22 patients with hydrocephalus were prospectively enrolled (Figure 1). Eight of the 22 patients with hydrocephalus had diagnoses other than iNPH and were enrolled in the control group (see their diagnosis in Figure 1). Ten of the 22 patients with hydrocephalus were diagnosed with probable or definite iNPH. Two patients with possible iNPH who neither exhibited DESH nor responded to spinal taps were excluded. Two patients without shunt responses were analyzed separately as shunt non-responders 1 and 2 (Figure 1). The clinical data of the participants are shown in Table 1. Age, sex, and disease duration in patients with PSP were not significantly different from those in the iNPH and control groups. The ratio of patients with vertical supranuclear gaze palsy or fall tendency in the PSP group was significantly higher than that in the iNPH (P = 0.0004, 0.0004, respectively) and control groups (P = 0.001, 0.0004, respectively). The iNPHGS gait domain and MDS-UPDRS part III scores were significantly higher in patients with PSP than in controls (P = 0.007 and 0.002, respectively). The imaging parameters at baseline are listed in Table 1. No significant differences in Evans index, callosal angle in the coronal plane at the posterior commissure, callosal angle at the splenium, or DESH score were observed between patients with PSP and iNPH or control. Three of the 11 patients with PSP had a high DESH score ≥ 5. The ratio of patients with DAT-SPECT abnormalities in patients with PSP was significantly higher than that in patients with iNPH (P < 0.0001) and control patients (P = 0.002).

**Figure 1.**
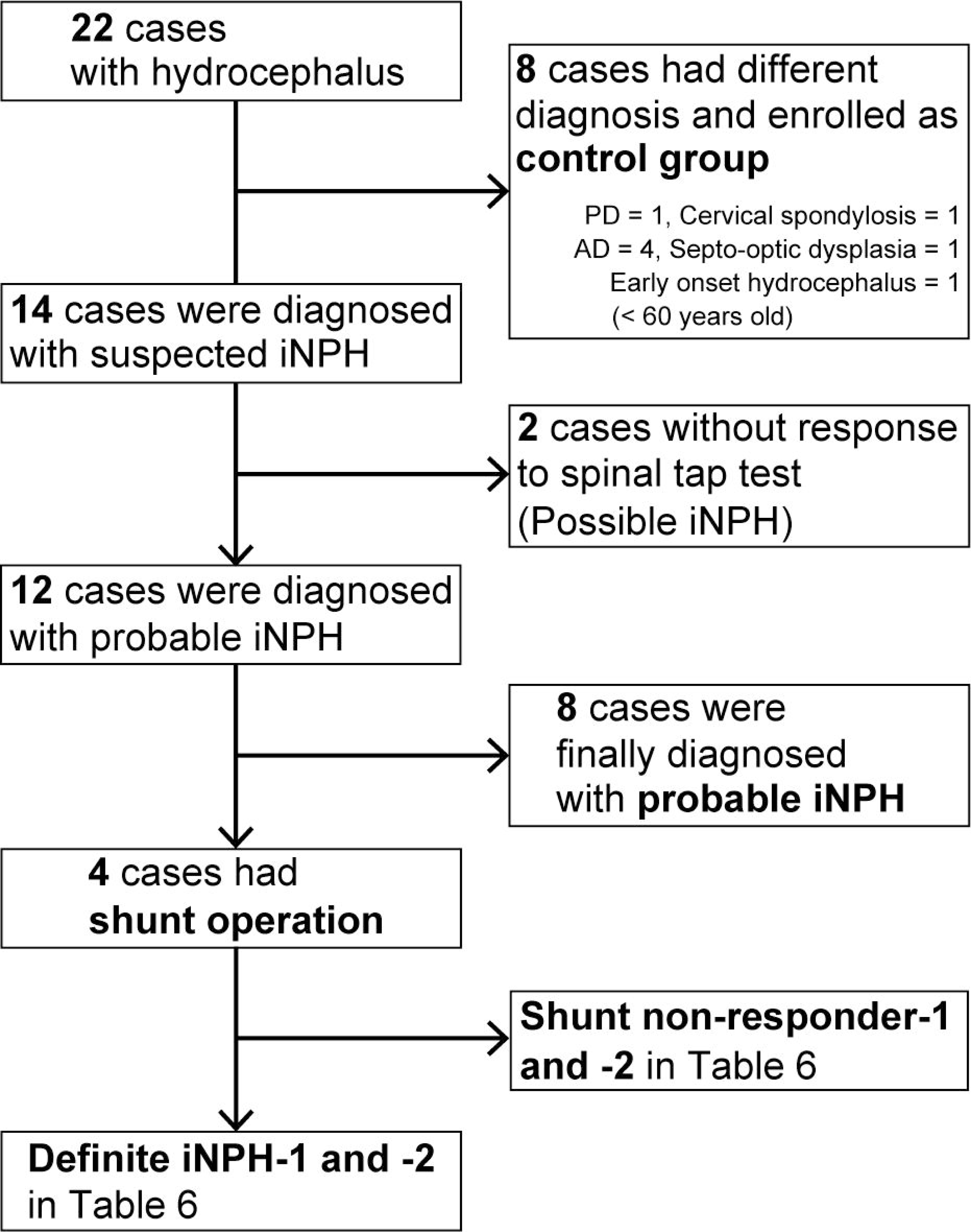
Flow diagram of the enrolled patients with hydrocephalus. PD, Parkinson’s disease; AD, Alzheimer’s disease; iNPH, idiopathic normal pressure hydrocephalus.

**Table 1.**
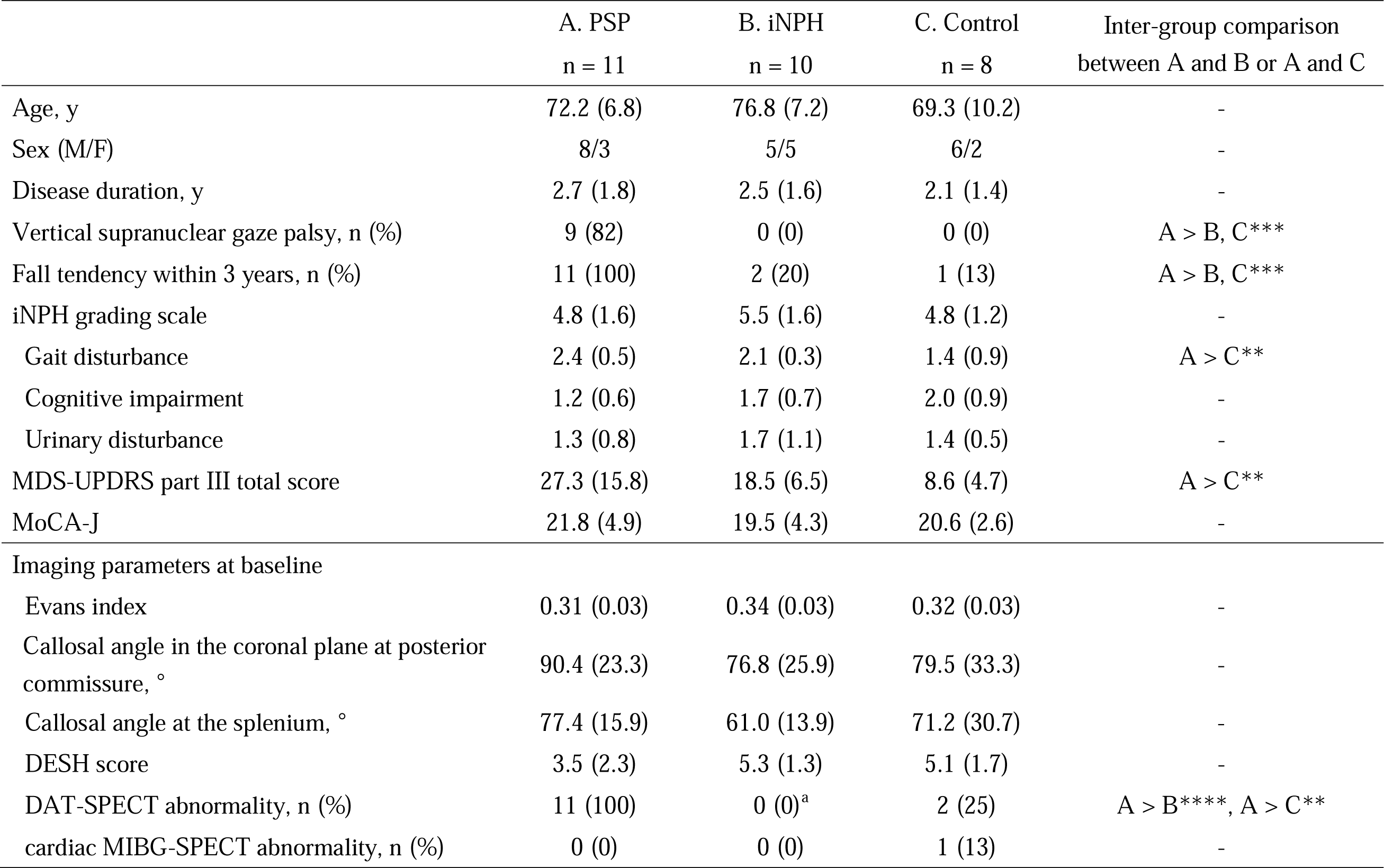
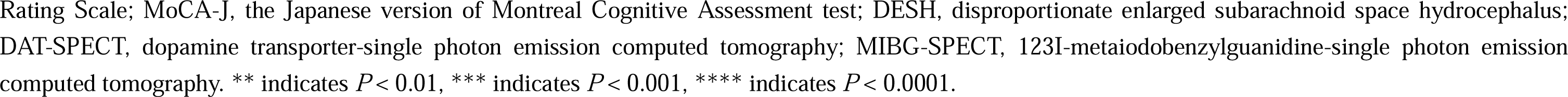
Characteristics of participants. Data are presented as mean (standard deviation) except sex, vertical supranuclear gaze palsy, fall tendency, DAT, and cardiac MIBG-SPECT. ^a^ n = 9. Patients with fall tendency are defined as those who had repeated unprovoked falls or the tendency to fall on the pull-test. PSP, progressive supranuclear palsy; iNPH, idiopathic normal pressure hydrocephalus; MDS-UPDRS, Movement Disorder Society Unified Parkinson’s Disease Rating Scale; MoCA-J, the Japanese version of Montreal Cognitive Assessment test; DESH, disproportionate enlarged subarachnoid space hydrocephalus; DAT-SPECT, dopamine transporter-single photon emission computed tomography; MIBG-SPECT, 123I-metaiodobenzylguanidine-single photon emission computed tomography. ** indicates *P* < 0.01, *** indicates *P* < 0.001, **** indicates *P* < 0.0001.

### Inter-trial comparison of the spinal tap/sham tap effect on gait function

Gait variables in the three groups at baseline, after the sham spinal tap, and after the spinal tap are shown in Table 2. For patients with PSP, the TUG total time at post-spinal tap was significantly shorter than that at baseline (P = 0.038). The dual-task stride velocity at post-spinal tap was significantly higher than at baseline (P = 0.028). There were no significant differences in the gait parameters between baseline and after the sham spinal tap. For patients with iNPH, the single-task stride length and velocity at the post-spinal tap were significantly higher than at baseline (P = 0.028 and 0.007, respectively). There were no significant differences in gait parameters between baseline and after the sham spinal tap or spinal tap in control patients.

**Table 2.**
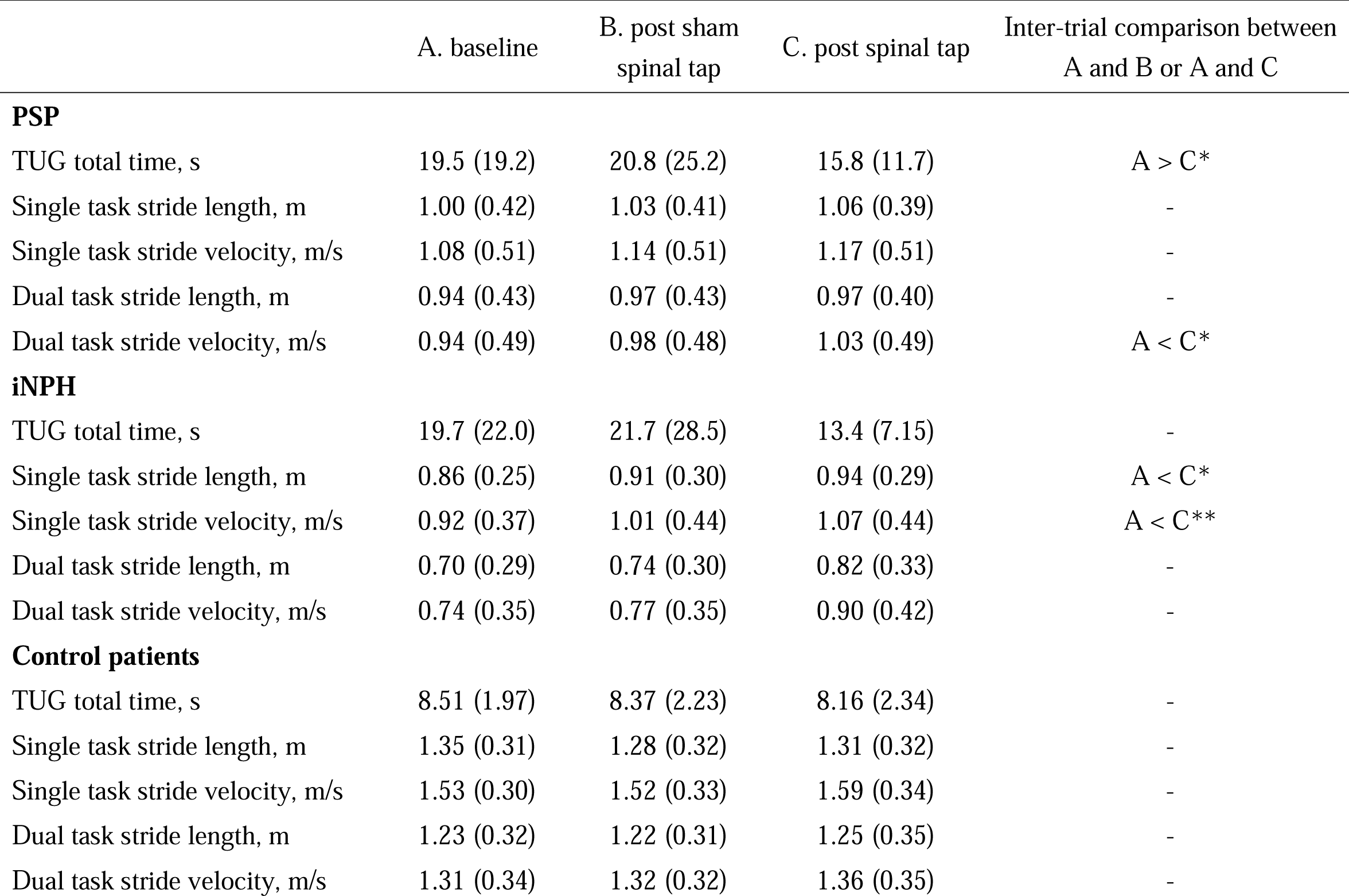
Gait variables before/after sham spinal tap and tap test Data are presented as mean (standard deviation). * indicates P < 0.05 and ** indicates P < 0.01 by the Friedman. test followed by Dunn’s test. See abbreviations in Table 1 legends.

### Inter-group comparison of responder ratio on gait function to the spinal tap/sham spinal tap

The ratios of responders to the spinal taps and sham spinal taps in each group are shown in Table 3. The proportion of patients with improvement in any gait parameter after the spinal tap in the PSP group (9 out of 11 patients, 82%) was significantly higher than that in the control group (2 out of 8 patients, 25%; P = 0.047), but it was not significantly different from that in iNPH group (9 out of 10 patients, 90%; P > 0.999). The proportion of patients with improvement in any gait parameter after the sham spinal tap in patients with PSP (8 out of 11 patients, 73%) was also significantly higher than that in control patients (1 out of 8 patients, 13%; P = 0.040), but it was not significantly different from that in patients with iNPH (6 out of 10 patients, 60%; P > 0.999). The ratio of patients with improvement in any gait parameter after the spinal tap, as compared to the better gait parameters at the baseline or post-sham spinal tap measurements, did not demonstrate a significant difference between patients with PSP (5 out of 11 patients, 45%), patients with iNPH (6 out of 10 patients, 60%; P > 0.999), or control patients (2 out of 8 patients, 25%; P > 0.999).

**Table 3.** The ratio of the responders showing at least 10% improvements in gait parameters after the spinal tap or the sham spinal tap from baseline. Data are presented as % (numbers of subjects). * indicates *P* <0.05. Data are presented as % (numbers of subjects). See abbreviations in. **Table 1 legends.**

|  | A. PSP<br>n = 11 | B. iNPH<br>n = 10 | C. Control<br>n = 8 | Inter-group comparison<br>between A and B or A and C |
| --- | --- | --- | --- | --- |
| **baseline vs. post spinal tap** |  |  |  |  |
| Gait parameters,<br>% (the number of responders) |  |  |  |  |
| TUG total time | 55% (6) | 50% (5) | 25% (2) | - |
| Single task stride length | 36% (4) | 40% (4) | 0% (0) | - |
| Single task stride velocity | 45% (5) | 70% (7) | 25% (2) | - |
| Dual task stride length | 40% (4) | 60% (6) | 13% (1) | - |
| Dual task stride velocity | 40% (4) | 70% (7) | 13% (1) | - |
| Any of the above gait parameters | 82% (9) | 90% (9) | 25% (2) | A > C* |
| **baseline vs. post sham spinal tap** |  |  |  |  |
| Gait parameters,<br>% (the number of responders) |  |  |  |  |
| TUG total time | 27% (3) | 30% (3) | 13% (1) | - |
| Single task stride length | 27% (3) | 10% (1) | 0% (0) | - |
| Single task stride velocity | 45% (5) | 60% (6) | 13% (1) | - |
| Dual task stride length | 30% (3) | 30% (3) | 0% (0) | - |
| Dual task stride velocity | 40% (4) | 40% (4) | 13% (1) | - |
| Any of the above gait parameters | 73% (8) | 60% (6) | 13% (1) | A > C* |

Within the patient with PSP cohort, the ratio of responders to gait function to the spinal tap and the sham spinal tap was compared between patients with PSP with a high DESH score ≥ 5 and patients with PSP with a low DESH score < 5 (Table 4). The ratio of patients with improvement in TUG total time after the spinal tap in patients with PSP with DESH score ≥ 5 (3 out of 3 patients, 100%) was significantly larger than that in patients with PSP with DESH score < 5 (1 out of 8 patients, 13%; P = 0.024).

**Table 4.** The comparison of the ratio of the responders between PSP patients with high DESH score and low DESH score Data are presented as % (numbers of subjects). See abbreviations in. **Table 1 legends.**

| | PSP with DESH score $\geq 5$ | PSP with DESH score $< 5$ | Fisher's exact test |
| --- | --- | --- | --- |
|  | n = 3 | n = 8 | P Value |
| **baseline vs. post spinal tap** |  |  |  |
| Gait parameters,<br>% (the number of responders) |  |  |  |
| TUG total time | 67% (2) | 50% (4) | - |
| Single task stride length | 100% (3) | 13% (1) | A > C* |
| Single task stride velocity | 67% (2) | 38% (3) | - |
| Dual task stride length | 33% (1) | 38% (3) | - |
| Dual task stride velocity | 33% (1) | 38% (3) | - |
| Any of the above gait parameters | 100% (3) | 75% (6) | - |
| **baseline vs. post sham spinal tap** |  |  |  |
| Gait parameters,<br>% (the number of responders) |  |  |  |
| TUG total time | 0% (0) | 38% (3) | - |
| Single task stride length | 33% (1) | 25% (2) | - |
| Single task stride velocity | 33% (1) | 50% (4) | - |
| Dual task stride length | 33% (1) | 25% (2) | - |
| Dual task stride velocity | 33% (1) | 38% (3) | - |
| Any of the above gait parameters | 33% (1) | 88% (7) | - |

### Inter-group comparison of responder ratio on cognitive function to the spinal tap

The cognitive functions of the tap responders were compared between the groups (Table 5). When at least 1.5 SD changes of the Z-score in two or more cognitive batteries were defined as responders, 5 out of 11 patients with PSP (45%) responded to the spinal tap, while 5 out of 10 patients with iNPH (50%) and 3 out of 8 control patients (38%) responded to the spinal tap. When at least 1.5 SD changes in the Z-score in three or more cognitive batteries were defined as responders, 1 out of 11 patients with PSP (9%) responded to the spinal tap, while 3 out of 10 patients with iNPH (30%), and 1 out of 8 control patients (13%) responded to the spinal tap. There was no significant difference between the groups in the ratio of patients with improved cognitive function with at least 2 batteries and at least 3 batteries.

**Table 5.**
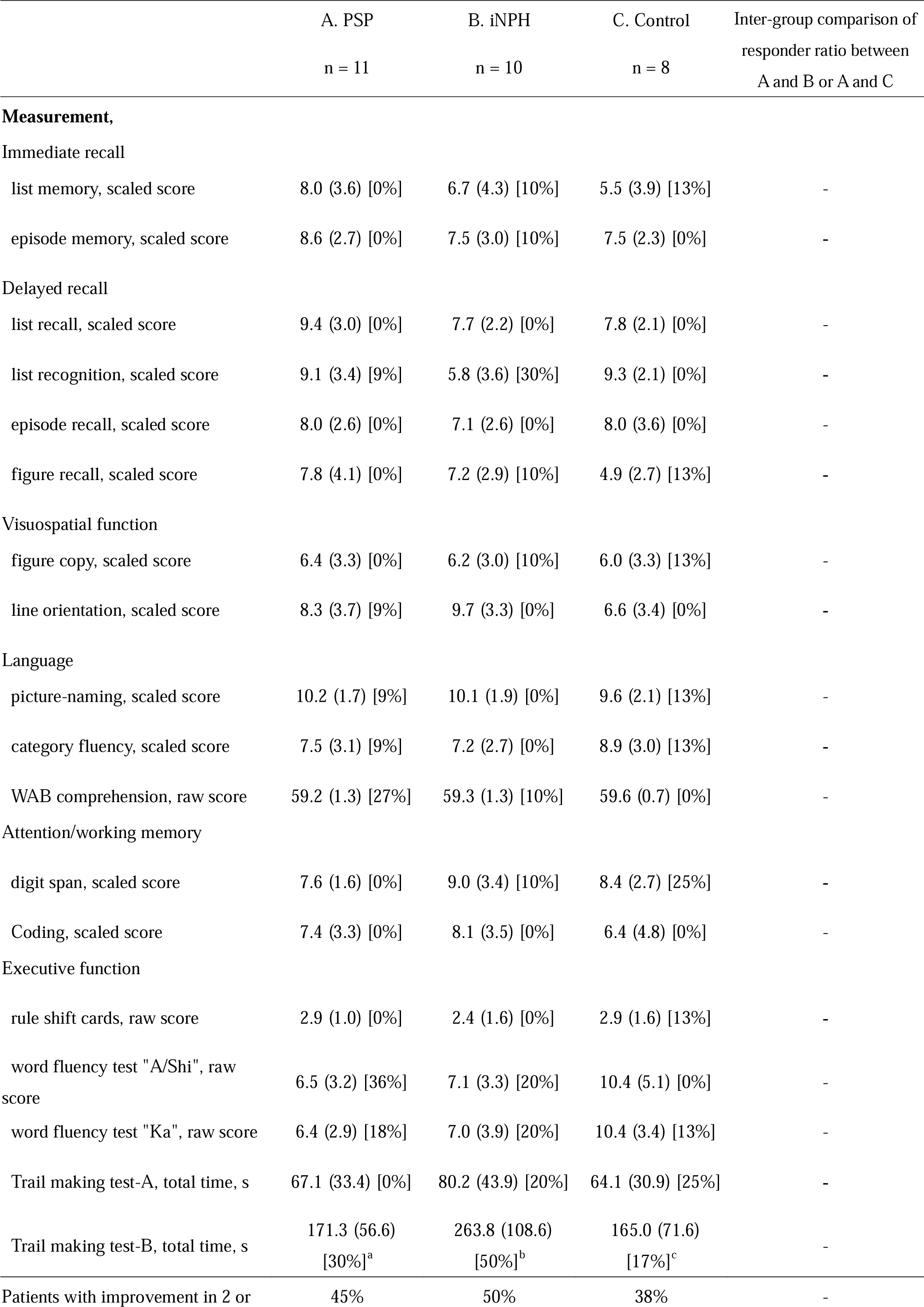
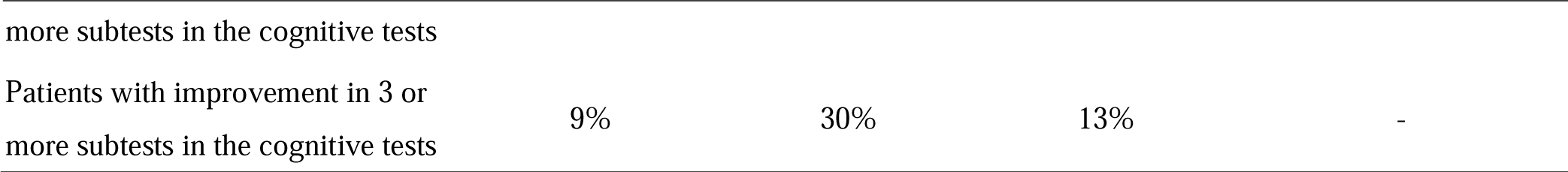
The ratio of the responders showing at least 1.5 SD improvements in cognitive tests after the spinal tap from baseline. Data are presented as the mean score (SD) at baseline [ratio of responders showing at least 1.5 SD improvements in the cognitive test after the spinal tap from baseline] ^a^ n = 10, ^b^ n = 8, ^c^ n = 6. See abbreviations in Table 1 legends.

### Analysis for patients who had shunt operation

One of the 11 patients with PSP (Case 1 in Figure 2, Table 6) and 4 of the 12 patients with probable iNPH (Figure 1) underwent a shunt operation. The response rates of the gait parameters after sham spinal tap, spinal tap, and shunt operations are shown in Table 6. One patient with PSP-RS responded better to the spinal tap than the sham spinal tap and shunt operation. Two patients with definite iNPH (definite iNPH-1 and definite iNPH-2 in Table 6) also showed improvement beyond the effect of the sham spinal tap.

**Figure 2.**
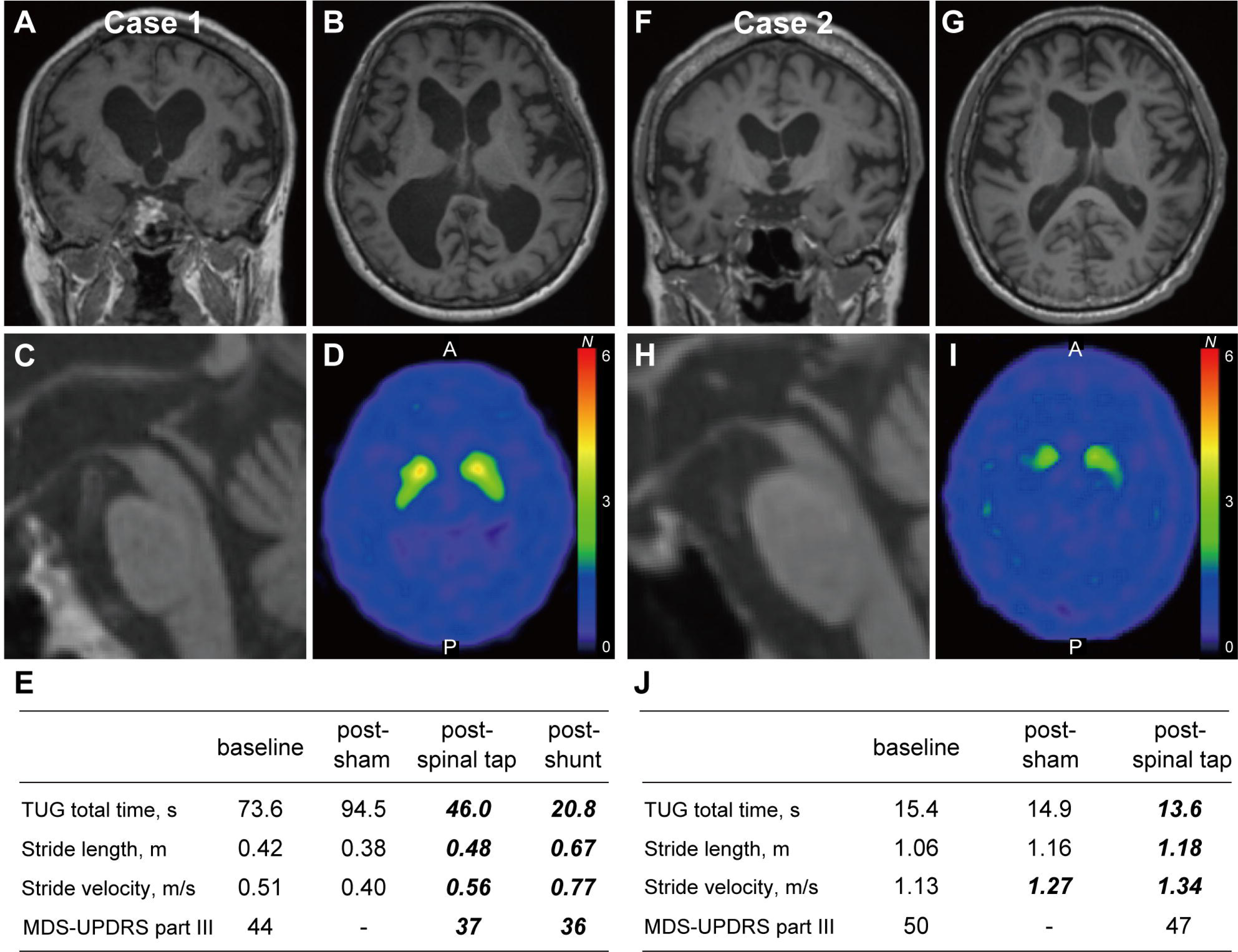
Representative PSP patients with spinal tap responsiveness. (A-C) T1-weighted images and (D) DAT-SPECT in a woman in her 70s with probable PSP-RS (Case 1). Right SBR was 1.79 and left SBR was 1.79. (E) Longitudinal gait/MDS-UPDRS part III assessments in Case 1. (F-H) T1-weighted images and (I) DAT-SPECT in a man in his 70s with probable PSP-RS (Case 2). Right SBR was 1.79 and left SBR was 2.49. (J) Longitudinal gait/MDS-UPDRS part III assessments in Case 2. TUG, timed up and go test; MDS-UPDRS, Movement Disorder Society Unified Parkinson’s Disease Rating Scale; SBR, Specific Binding Ratio. Improvements by more than 10% from the baseline are shown in bold italics.

**Table 6.** The response ratio of gait variables compared with baseline for individuals who had shunt operation Data are presented as the ratio of improvements in the gait variables compared to those from baseline/post-sham tap. Improvements with more than 10% from baseline/post-sham tap were shown in bold italics. PSP-RS, progressive supranuclear palsy-Richardson syndrome.

| Participants who had shunt operation | baseline vs. post-sham tap | baseline vs. post-spinal tap | post-sham tap vs. post-spinal tap | post-sham tap vs. post-shunt |
| --- | --- | --- | --- | --- |
| PSP-RS (Case 1 in Figure 2) |  |  |  |  |
| TUG total time | -28.4% | **37.5%** | **51.3%** | **78.0%** |
| Single task stride length | -9.5% | **14.3%** | **26.3%** | **76.3%** |
| Single task stride velocity | -21.6% | 9.8% | **40.0%** | **92.5%** |
| Definite iNPH-1 |  |  |  |  |
| TUG total time | **23.8%** | **33.6%** | **12.9%** | **23.8%** |
| Single task stride length | 4.9% | **16.0%** | **10.6%** | **24.7%** |
| Single task stride velocity | 9.6% | **32.9%** | **21.3%** | **35.0%** |
| Definite iNPH-2 |  |  |  |  |
| TUG total time | -26.2% | **19.3%** | **36.0%** | 5.1% |
| Single task stride length | -1.3% | 6.7% | 8.1% | **12.2%** |
| Single task stride velocity | -6.2% | **35.4%** | **44.3%** | **39.3%** |
| Shunt non-responder-1 |  |  |  |  |
| TUG total time | 3.1% | **10.2%** | 7.4% | -3.2% |
| Single task stride length | **24.4%** | **24.4%** | 0% | -4.9% |
| Single task stride velocity | **30.3%** | **31.5%** | 0.9% | -9.5% |
| Shunt non-responder-2 |  |  |  |  |
| TUG total time | 5.7% | **13.2%** | 7.9% | -12.1% |
| Single task stride length | 2.4% | **12.2%** | 9.5% | -9.5% |
| Single task stride velocity | **13.5%** | **19.8%** | 5.5% | -18.3% |

However, two shunt non-responders (shunt non-responder-1 and shunt non-responder-2) did not show improvement in gait parameters beyond the effect of the sham spinal tap after the spinal tap.

Two representative patients with PSP-RS who responded to spinal taps are shown in Figure 2. Case 1 is a woman in her 70s presented with the typical symptoms of PSP-RS, including supranuclear vertical gaze palsy and postural instability (Figure 2A-E). MRI showed slight atrophy of the midbrain, ventricular enlargement, a dilated Sylvian fissure, and a high tight convexity, suggesting the presence of iNPH-like MRI features (Figure 2A-C; EI: 0.34; DESH score: 5; CA at the posterior commissure: 63.3°; CA at the splenium: 58.8°). Although DAT-SPECT showed dopaminergic denervation (Figure 2D), cardiac MIBG-SPECT results were normal. Her gait parameters and MDS-UPDRS part III scores improved after spinal tap and shunt operations (Figure 2E). In case 2, a man in his 70s presented with progressive gait difficulties and frequent falls (Figure 2F-J). Neurological examination revealed supranuclear vertical gaze palsy and Parkinsonism with marked postural impairment. Brain MRI revealed midbrain atrophy and dilatation of the lateral ventricles (Figure 2F-H; EI: 0.33; DESH score: 4; CA at the posterior commissure: 72.4°; CA at the splenium: 83.1°). While DAT-SPECT showed dopaminergic denervation (Figure 2I), cardiac MIBG-SPECT findings were normal. All gait parameters were improved by 10% or more after the spinal tap compared with those at baseline, but their improvement was less than 10% compared with those after the post-sham spinal tap (Figure 2J). The MDS-UPDRS part III scores did not change after the spinal tap.

## Discussion

This prospective study is the first to demonstrate the favorable effects of spinal tap/sham spinal tap in patients with PSP. This study has three main findings. First, most patients with PSP gait parameters responded positively to spinal taps. Second, the gait parameters of most patients with PSP also responded positively to the sham spinal tap, which was also observed in patients with iNPH. Third, one patient with PSP, in addition to the patients with iNPH, exhibited favorable responses to the shunt operation, and all showed responses to the spinal tap beyond the placebo effect before the shunt operation.

In our study, nine of the 11 patients with PSP showed notable improvements in at least one gait parameter after the spinal tap. The ratio of responders to the spinal taps in patients with PSP was significantly larger than that in control patients. Furthermore, within the patient with PSP cohort, certain gait parameters showed significant improvement following the spinal taps compared to their baseline values (Table 2). Notably, patients with PSP with a high DESH score (≥ 5), which serves as an indicator of iNPH-like MRI features,^4^ tended to show a higher response rate to the spinal tap in terms of TUG total time than patients with PSP with a low DESH score (< 5) (Table 4). Based on these results, it is possible that gait disturbances in certain patients with PSP, particularly those with a high DESH score, tend to respond favorably to spinal taps.

It should be noted that the improvement in gait parameters observed in patients with PSP may also be attributed to the placebo effect. In our study, eight of the 11 patients with PSP exhibited a response to the sham spinal tap, and the ratio of the sham spinal tap responders was significantly greater than that of control patients, but not greater than that of patients with iNPH (Table 3). In Figure 2, Case 2 did not show improvement in gait parameters exceeding 10% after the spinal tap compared to the post-sham tap, and he did not undergo a shunt operation. In contrast, Case 1, who improved gait parameters beyond the placebo effect after the spinal tap, responded to the shunt operation. This patient presented with iNPH-like MRI features with a high DESH score, which we previously proposed as PSP-H.^4^ Patients with PSP-H may exhibit iNPH-like clinical presentations, spinal tap/shunt responsiveness, as well as iNPH-like MRI features. Moreover, it is conceivable that the shunt operation may effectively alleviate gait disabilities even in patients with PSP-H if they exhibit a response to the spinal tap surpassing the placebo effect.

Several previous studies have shown that the placebo effect may be associated with improved clinical symptoms in patients with iNPH following a spinal tap or shunt operation.^11,13^ In our study, although 92% of the patients with iNPH responded to the spinal tap, 67% responded to the sham spinal tap, indicating that the placebo effect, at least in part, contributed to the improvement in gait performance among some patients with iNPH. Furthermore, the sham spinal tap may be useful in preventing overestimating the effects of the spinal tap. In our study, four patients with iNPH who underwent shunt operations highlighted the usefulness of the sham spinal tap (Table 6). The two patients who responded to the shunt operation (definite iNPH-1 and definite iNPH-2) also responded to the spinal tap beyond the placebo effect. However, the other two patients who did not respond to the shunt operation (shunt non-responder-1 and shunt non-responder-2) did not respond to the spinal tap beyond the placebo effect. Therefore, a sham spinal tap can help to avoid unnecessary shunt operations.

The definition of “responder” may have an impact on the results of our study. We used a cut-off value of 10% improvement in the gait parameter to classify responders after each test compared to baseline.^29^ A recent Japanese guideline on iNPH suggested that a cut-off value of 10% could result in more false positives for the spinal tap in patients with iNPH. According to Yamada et al., a reduction in TUG total time by more than 5 s was reported to be more specific for identifying those who responded to the shunt operation. However, it is not sensitive enough to detect shunt responders with mild gait disturbance.^38^ Therefore, the appropriate criteria for defining a spinal tap responder who will respond positively to shunt operations have not yet been established. On the contrary, a review study conducted by Mihalj et al. demonstrated that the sensitivity of the spinal tap in predicting shunt response was only 58%,^12^ suggesting that a negative response to the spinal tap does not necessarily exclude these patients from being suitable candidates for the shunt operation. We used a cut-off value of 10% improvement for multiple gait measures in order to supplement this low sensitivity of the spinal tap.

Many reports have assessed cognitive function in patients with iNPH before and after the spinal tap using the MMSE total score^39–42^ and have suggested that the MMSE total score improved following the spinal tap.^39^ However, the MMSE can be influenced by a learning effect^43^ and may only evaluate limited cognitive domains. In this study, we employed the RBANS, designed to avoid the learning effect, using two independent sets of batteries. We conducted a comprehensive neurocognitive assessment by thoroughly evaluating all cognitive domains of the participants. Unlike the gait function, there was no significant difference in the ratio of tap responders regarding cognitive function among the groups. Since the criteria to define cognitive function responders to the spinal tap were not established, we employed two different cut-offs to ascertain the inter-group differences: (a) at least 1.5 SD improvement in two or more subtests: 45% of patients with PSP, 50% of patients with iNPH, and 38% of control patients responded, and (b) at least 1.5 SD improvement in three or more subtests: 9% of patients with PSP, 25% of patients with iNPH, and 13% of control patients responded (Table 4). In conclusion, cognitive function may be, at least in part, improved after the spinal tap test in patients with PSP, but this remains to be confirmed.

Given that iNPH is thought to impair cerebrospinal fluid circulation, it is reasonable to postulate that some patients with PSP may also have compromised cerebrospinal fluid circulation, resulting in iNPH-like MRI features, spinal tap responsiveness, or both. Aquaporin-4 (AQP4) water channels, which are predominantly localized in the glial cell membrane, particularly in the perivascular astroglial endfeet, play an important role in cerebrospinal fluid circulation. Previous histopathological studies have reported that the increasing severity of astrogliosis is associated with altered expression, impaired polarization, or both of AQP4 at the astrocytic perivascular endfeet in iNPH,^44^ as well as in other diseases/conditions.^45,46^ In contrast, previous pathological studies in patients with PSP^47,48^ have revealed a close relationship between tau pathology and astrogliosis. Based on these collective findings, we hypothesized that tau-induced astrogliosis leads to decreased AQP4 polarization at the perivascular astroglial endfeet in patients with PSP, thereby impairing cerebrospinal fluid circulation and manifesting iNPH-like phenotypes.

Our study had several limitations. First, it enrolled a limited number of patients, particularly those who underwent shunt surgery. A larger prospective study that enrolls more patients with PSP-H and definite iNPH patients is necessary to validate our findings. Second, all patients were diagnosed clinically, and they were not pathologically confirmed. However, in order to ascertain the pathological background, we conducted DAT-SPECT and cardiac MIBG-SPECT in all patients with PSP and 11 of the 12 patients with iNPH, supporting our diagnosis. Moreover, patients with PSP-RS who comprise the majority of our study cohort showed higher clinicopathological correlations,^49^ and therefore we expect that the diagnosis of PSP is reasonably valid. Finally, it is noteworthy that our control patients had diverse etiologies, and their responses to spinal taps were unknown. However, we could not include healthy elderly individuals in the spinal tap test because of ethical concerns.

In conclusion, patients with PSP, particularly those with a high DESH score, often demonstrate amelioration of their gait impairments following a spinal tap, and even after a shunt operation, similar to patients with iNPH. We propose the classification of PSP patients with iNPH-like MRI/clinical features as PSP-H. Future larger-scale studies are needed to characterize better the clinical and pathological features of PSP-H. Moreover, our thorough evaluation revealed that patients with both PSP and iNPH often manifest a placebo effect during gait assessment, which can be evaluated by implementing a sham spinal tap. Comparing the response to a spinal tap with that of a sham spinal tap may help accurately identify patients with PSP or iNPH who definitively respond to the shunt operation.

## Acknowledgments

The authors thank Taiki Matsubayashi, MD and Satoko Kina, MD for collecting gait data, Nobuo Sanjo, MD, PhD, Yui Nosaki, Miki Yoshitake and Meiko Hirai for collecting neuropsychological data, and the clinical staffs in our hospitals for taking care of patients.

## Author contributions

MO, TH, and TY contributed to the conception and design of the study; MO, TH, QC, KS, KH, and MM contributed to the acquisition and analysis of data; MO and TH contributed to drafting the text or preparing the figures.

## Data availability

Investigators may request access to anonymized data that was used in this study. Prior to using the data, proposals must be approved by the Institutional Review Boards at Tokyo Medical and Dental University Hospital.

